# Effect of experimentally introduced interaural frequency mismatch on sentence recognition in bilateral cochlear-implant listeners

**DOI:** 10.1101/2023.01.06.23284274

**Authors:** Miranda Cleary, Kristina DeRoy Milvae, Nicole Nguyen, Joshua G. W. Bernstein, Matthew J. Goupell

**Author notes:** Corresponding Author: Miranda Cleary, Department of Hearing and Speech Sciences, University of Maryland, College Park, MD 20742.

## Abstract

Bilateral cochlear-implant users experience interaural frequency mismatch because of asymmetries in array insertion and frequency-to-electrode assignment. To explore the acute perceptual consequences of such mismatch, sentence recognition in quiet was measured in nine bilateral cochlear-implant listeners as frequency allocations in the poorer ear were shifted by ±1.5, ±3 and ±4.5 mm using experimental programs. Shifts in frequency allocation >3 mm were found to reduce bilateral sentence scores below those for the better ear alone, suggesting that the poorer ear interfered with better-ear perception. This was not a result of fewer active channels; deactivating electrodes without frequency shifting had minimal effect.

## 1. INTRODUCTION

Bilateral cochlear implants (BI-CIs) offer advantages over unilateral cochlear implants (CIs) for sound localization and speech understanding in noise (Smulders *et al*., 2016). While having two CIs is usually beneficial, under challenging conditions such as multiple competing talkers, some BI-CI users perceive speech more poorly with bilateral inputs than with a unilateral input (Goupell *et al*., 2018a). Such results have been described as a form of contralateral interference. Analogous effects involving contralateral interference have been discussed as also affecting 10-20% of older bilateral hearing-aid users (Jerger and Silverman, 2018).

Contralateral interference could be related to interaural asymmetries in electrode location or device programming. Because of the cochlea’s tightly coiled structure, CI electrode arrays are often inserted only approximately one full basal turn of the cochlea relative to its full 2.5–2.75 turns. The array then delivers information about input frequencies between 0.2–8 kHz to locations tuned to higher frequencies in acoustic hearing, resulting in tonotopic frequency-to-place mismatch (Landsberger *et al*., 2015). Additionally, surgical or anatomical considerations sometimes result in different insertion depths, such that one-third of BI-CI users are estimated to have at least three electrode pairs with >3 mm of interaural frequency mismatch (Goupell *et al*., 2022). Asymmetric clinical deactivation of electrodes, due to facial nerve stimulation for example, can also introduce interaural frequency mismatch because the full range of input frequencies is typically reallocated to remaining active electrodes, irrespective of the allocation on the opposite side.

Interaural frequency mismatch reduces bilateral word recognition scores in normal-hearing individuals listening to vocoder CI simulations when compared with conditions with no mismatch (Yoon *et al*., 2011; van Besouw *et al*., 2013; Fitzgerald *et al*., 2017; Goupell *et al*., 2018b). Yoon *et al*. (2011) additionally found that a 3-mm interaural mismatch—simulating a shallow insertion on one side and typical insertion on the other—reduced bilateral scores in quiet to below unilateral performance with the typical insertion simulation. Thus, the distorted speech in the more shifted ear interfered with perception, rather than providing supplemental information.

These prior vocoder studies are suggestive of a relationship between interaural frequency mismatch and contralateral interference, but they did not involve actual BI-CI listeners. Although it is not practical or ethically feasible for researchers to physically relocate existing electrode arrays to manipulate interaural mismatch, it is possible to change interaural frequency alignment through adjustments to clinical programs. The present study therefore assessed if bilateral sentence scores were influenced by manipulating frequency allocations in the poorer ear of BI-CI listeners, with scores for the unshifted better ear alone serving as the baseline. If a shifted signal in the poorer ear could be ignored, we hypothesized that bilateral scores would be no worse than for the better ear alone. This preliminary study tested sentence recognition in quiet (in preparation for future planned testing in noise), expecting that interference effects were unlikely to be seen under such favorable listening conditions. Surprisingly, interference effects were observed, increasing with degree of interaural mismatch.

## 2. METHOD

### 2.1 Participants

Nine post-lingually deafened adult BI-CI users participated (mean age: 67 yrs, range: 34– 82 yrs). Each participant had >6 months of bilateral experience with Cochlear-brand CIs, which are intended to have 22 active intracochlear electrodes, although in some cases individual electrodes were clinically deactivated (Table 1). Participants used either the CP910, 920, 950, or 1000 as their everyday CI speech processors. Additional details are shown in Table 1. All participants passed a brief cognitive screening, scoring ≥ 26 on the MoCA (Nasreddine *et al*., 2005). Procedures were approved by the Institutional Review Board at University of Maryland– College Park and participants provided informed consent.

**TABLE 1.** Device and program details for individual participants.

| Subject Code | Re-programmed Poorer Ear | Left Internal | Right Internal | Total # of Clinically Activated Electrodes | | Deactivated Electrodes | | # of Electrodes In Baseline $\Delta 0$ Program |
| --- | --- | --- | --- | --- | --- | --- | --- | --- |
|  |  |  |  | Left | Right | Left | Right |  |
| P01 | LEFT | Freedom (CI24RE) | Freedom (CI24RE) | 20 | 20 | 1,2 | 1,2 | 10 |
| P02 | LEFT | Freedom (CI24RE) | Freedom (CI24RE) | 22 | 22 | n/a | n/a | 11 |
| P03 | LEFT | CI512 | Freedom (CI24RE) | 22 | 22 | n/a | n/a | 11 |
| P04 | RIGHT | CI422 | CI24R (CS) Nucleus 24 Contour | 20 | 19 | 1,2 | 1,2,3 | 10 |
| P05 | RIGHT | Freedom (CI24RE) | Freedom (CI24RE) | 22 | 19 | n/a | 8,16,21 | 11 |
| P06 | LEFT | CI512 | Freedom (CI24RE) | 22 | 22 | n/a | n/a | 11 |
| P07 | RIGHT | CI512 | Freedom (CI24RE) | 21 | 21 | 1 | 1 | 11 |
| P08 | LEFT | Freedom (CI24RE) | Freedom (CI24RE) | 21 | 21 | 1 | 1 | 11 |
| P09 | RIGHT | CI24R (CS) Nucleus 24 Contour | Freedom (CI24RE) | 22 | 22 | n/a | n/a | 11 |

### 2.2 Procedure

#### 2.2.1 Stimuli

Institute of Electrical and Electronics Engineers (IEEE) Harvard sentences (IEEE, 1969) recorded by a male talker were used to measure sentence recognition. Each contained five different keywords.

#### 2.2.2 Experimental Programs

A spectral-peak-picking CI sound processing strategy that activates subsets of electrodes per stimulation cycle can introduce differences in the signals across the ears at a given point in time (Gray *et al*., 2021). To avoid this potential confound, clinical CI programming software (Custom Sound 5.2 or 6.2, Cochlear Ltd., Sydney, Australia) was used to convert each participant’s clinical spectral-peak-picking programs to experimental programs that adopted a continuous interleaved sampling strategy (Wilson *et al*., 1991) and frequency-aligned settings. These programs used every other electrode and set the number of maxima equal to the number of active electrodes, usually 11 electrodes covering the clinical frequency range of 188–7938 Hz [Fig. 1(A)].

**Fig 1.**
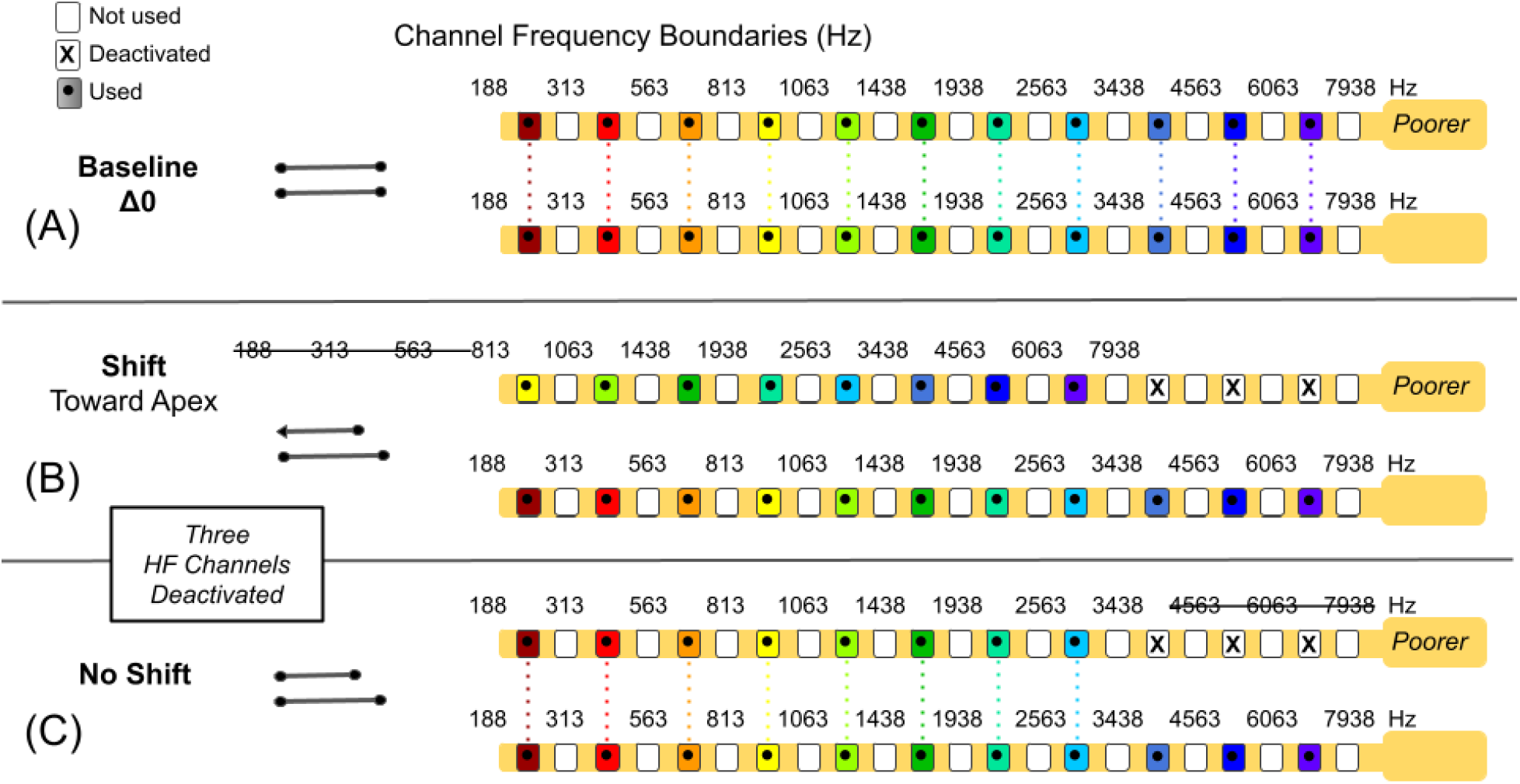
Example bilateral conditions. (A) Frequency-aligned programs with 11 active even-numbered channels were used in the Baseline Δ0 condition. In the experimental conditions, 1, 2 or 3 (this example) electrodes were further deactivated in the poorer ear at the high-frequency (HF, this example) or low-frequency end of the array. (B) In each Shift condition, frequency allocations were moved towards the apex or base by an equivalent number of electrodes, and unallocated frequencies at the end of the array were removed. (C) In No-Shift control conditions, the same electrodes were deactivated but with no frequency shift.

For any participant with clinically deactivated electrodes (Table 1), the corresponding electrodes in the opposite array were also deactivated. These programs with matched electrodes and frequency allocations served as the baseline “Δ0” condition. Only these baseline settings were used in the better ear.

The poorer-ear experimental programs were created by further deactivating 1, 2, or 3 electrodes at the low-frequency (LF) or high-frequency (HF) end of the array. These usually corresponded at the HF end to electrode 2 (1HF), electrodes 2 and 4 (2HF), or electrodes 2, 4, and 6 (3HF). At the LF end, these usually corresponded to electrode 22 (1LF), electrodes 22 and 20 (2LF), or electrodes 22, 20, and 18 (3LF).

The primary manipulation of interest was frequency shift. In the Shift conditions [Fig. 1(B)], frequencies previously assigned in the baseline condition to now-deactivated electrodes were shifted to the nearest active electrodes. These electrodes were 2, 4, or 6 electrodes higher or lower, corresponding to shifts of approximately 1.5, 3, or 4.5 mm along the cochlea. Frequency allocations for all other electrodes were also shifted by the same number of electrodes and displaced frequency bands eliminated at the opposite end (i.e., no frequency compression was applied). For BI-CI listeners, the majority of whom have relatively well-matched arrays (Goupell *et al*., 2022), shifted allocations mimic interaural frequency mismatch. An additional No-Shift condition [Fig. 1(C)] controlled for the loss of frequency information separate from the effects of shift, with frequencies previously assigned to deactivated electrodes simply removed from the signal. Only the 3LF and 3HF No-Shift conditions were included because pilot testing showed negligible effects with fewer deactivated channels. A total of 9 different programs (1 Baseline, 6 Shift, 2 No-Shift) were therefore created for the poorer ear.

The better ear was tested alone (1 unilateral condition) or paired with each poorer-ear experimental program (9 bilateral conditions). Additionally, the poorer ear was tested alone with each experimental program (9 unilateral conditions), for a total of 19 conditions. The rationale for reprogramming only the poorer ear was to avoid making changes to the better ear upon which BI-CI users often rely more heavily.

On the first day of testing, a research audiologist performed basic device diagnostics (i.e., they checked device impedances and voltage compliances). They then fit the participant with the baseline experimental program first in the better ear and then in the poorer ear. Levels were mostly left unchanged from those in the participant’s clinical programs. For participant P08, one electrode’s comfort “C” level was reduced to not exceed the compliance limit. Experimental programs were stored on laboratory clinical sound processors (N6, Cochlear Ltd.) dedicated to research purposes. Preprocessing features intended to improve speech perception in sound-field listening conditions (noise-reduction, audibility boosting, and SCAN options) were disabled. The default mostly omnidirectional Standard microphone mode was used. Volume and sensitivity were left at each participant’s preferred levels, usually the manufacture defaults of 6 and 12, respectively.

#### 2.2.3 Stimulus Presentation

Stimuli were presented via circumaural headphones (Sennheiser HD650, Hanover, Germany) placed around and over behind-the-ear CI sound processors (Goupell *et al*., 2018a). During unilateral testing, the processor on the opposite side was removed or muted.

Testing occurred in a sound attenuated testing booth (International Acoustics Co., Bronx, NY). The stimulus level began at 65 dB-A, and the volume was then adjusted to a comfortably loud and interaurally balanced listening level. Adjustments were made primarily to the volume in the poorer ear (better-ear exceptions: P01: upward, P08 and P05: downward) and changes were limited to ±5 dB for a maximum of 70 dB-A. Level adjustment was done exclusively at the beginning of the session using the baseline programs and a recorded sentence with the same average root-mean-square amplitude as the test stimuli. Tasks were run using software developed for MATLAB (Mathworks, Natick, MA) running on a personal computer.

#### 2.2.4 Measurement of Sentence Recognition

On each trial, one sentence was presented to either a single CI or both CIs. The sentence was selected at random from the 720 IEEE sentences. The selection was made without replacement until the available sentences were exhausted for a given participant, at which point the selection began anew from the full set. Participants repeated the sentence aloud, while an experimenter scored which of the five keywords were correctly repeated. No feedback was provided.

For each of the 19 conditions, 60 sentences were tested in sets of 20 trials, one set per main testing block. For each of the three main testing blocks, a different randomization of the 9 poorer-ear experimental programs was generated for each participant. The bilateral and unilateral conditions were counterbalanced as fully as possible across the three main testing blocks but were tested in groups of 4 experimental programs (one per sound-processor slot) to minimize the number of times the programs were changed and the CI processors and headphones were repositioned. Participants completed two main testing blocks on the first day of testing. On the second day, participants completed the third main testing block (except P08 who only completed two blocks), then took a break to converse and socialize with the study team while wearing the 3HF Shift program in the poorer ear and the baseline program in the better ear. (The 3HF Shift condition in pilot testing appeared to be the most difficult.) After this additional experience, 60 more sentences were tested using the 3HF Shift program. We also collected, either after all experimental testing or on a different day, sentence scores in quiet using the participant’s own processors and settings. Thus, the testing required a total of at least 1440 sentences per participant. Although sentences were repeated, this repetition was randomly distributed across conditions.

### 2.3 Analysis

Linear mixed effect models (LMEMs) assessed the within-subject effects of experimental programs and number of inputs/ears. Each model included random by-participant-varying intercepts and used the Satterthwaite *df* approximation to calculate denominators for *t* and *F* statistics. An α level of .05 was assumed.

## 3. RESULTS

Sentence recognition scores are plotted as a function of the number of deactivated channels, shift condition, and ear of presentation in Fig. 2 (individuals) and Fig. 3 (group means). Most participants incurred only a small drop in bilateral performance for the baseline experimental programs (filled green square at Δ0) compared to their everyday clinical programs (text labels in Fig. 2), despite the experimental program’s use of novel every-other electrode frequency allocations and more active channels. The average decrease of 7.1% (range: -3.7% to 30.3%; everyday programs, *M*=89.9%; experimental Δ0 programs, *M*=82.8%) did not reach significance [*t*(9)=2.13, *p*=.06].

**Fig 2.**
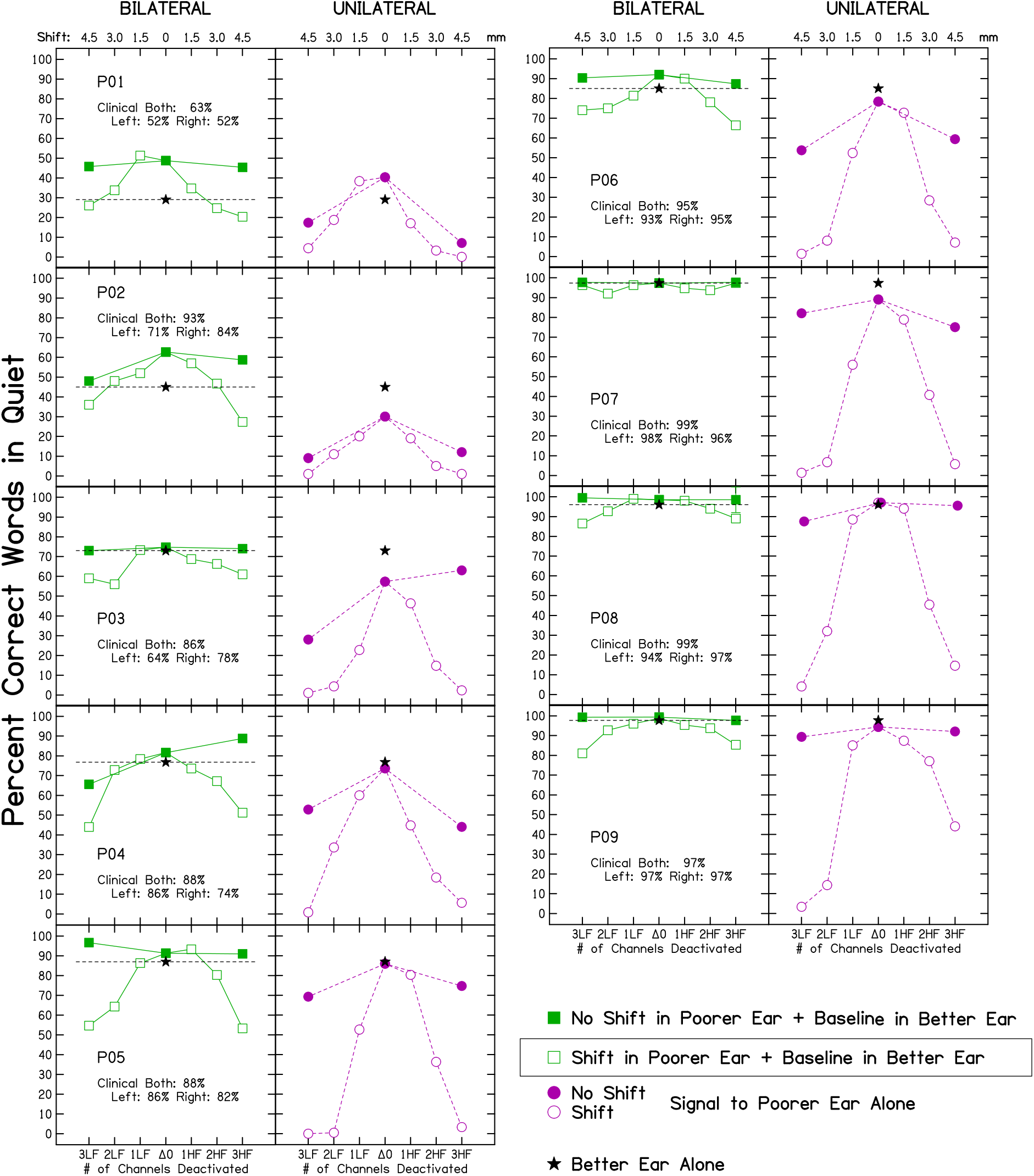
Sentence recognition scores for individual participants. Scores using their clinical programs are shown as text in individual panels. The box in the legend identifies the bilateral test condition of primary interest.

**Fig 3.**
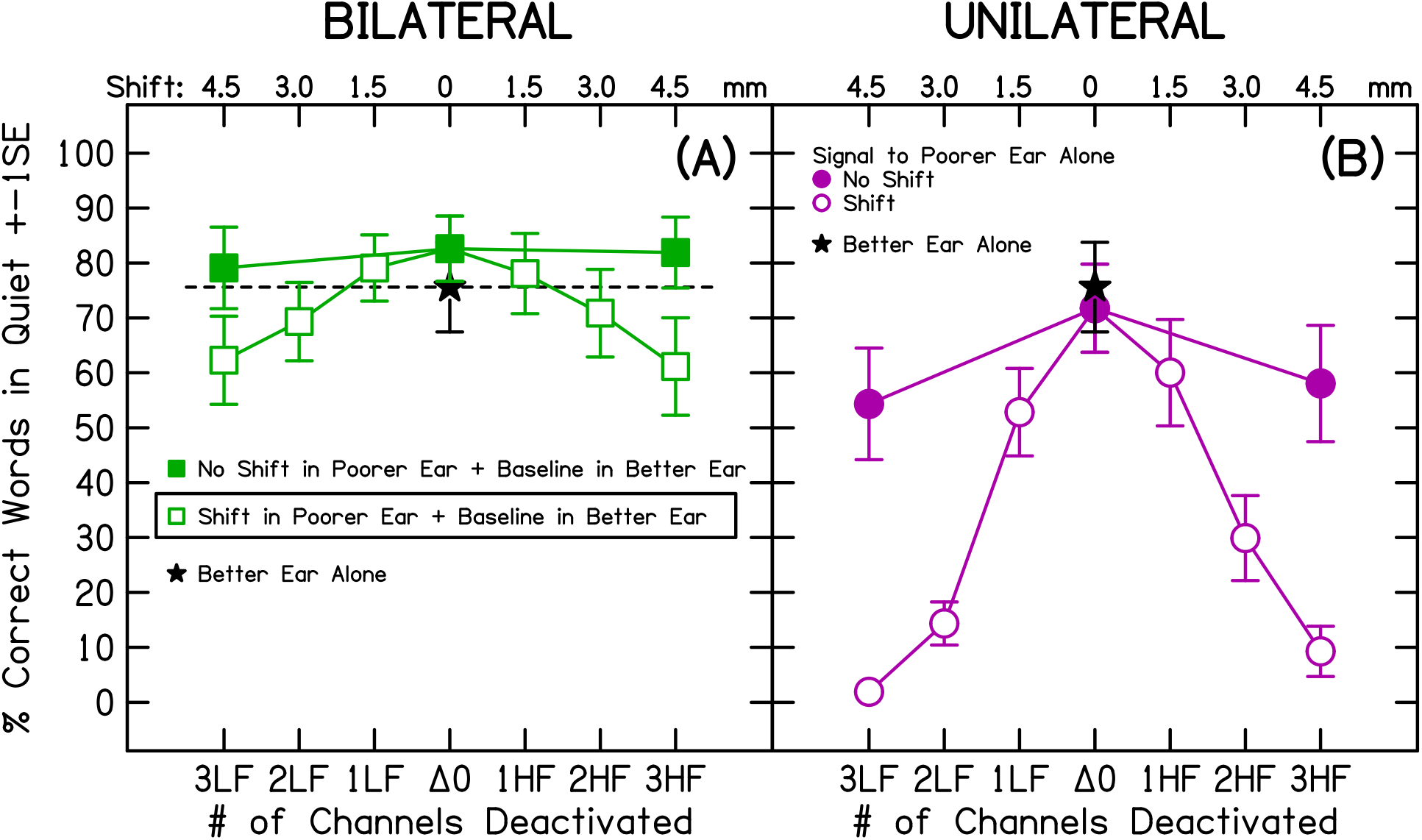
Average sentence recognition scores for bilateral (squares, panel A) and unilateral (poorer ear, circles, panel B) conditions. Filled symbols indicate control conditions with no frequency shift. Open symbols show shifted conditions. Unilateral scores for the better ear alone are shown by star symbols. The box in the legend identifies the bilateral test condition of primary interest.

The flat trajectory for the No-Shift control conditions (filled squares at 3LF and 3HF) in Fig. 3(A) shows that eliminating three channels of information at either end of the frequency range in the poorer ear without shifting did not change bilateral performance [*F*(2,18)=1.33, *p*=.29]. Frequency shifts in the poorer ear, however, had a clear detrimental effect when listening bilaterally (open squares). Scores decreased for eight of nine participants (all but P07, Fig. 2).

The average decrease for a three-channel shift was approximately 20% [3LF=20.9%, 3HF=21.6%]. A LMEM analysis with experimental program as a categorical variable and Δ0 as the reference level showed a main effect of experimental program [*F*(6,54)=17.61, *p*<.001]. Nine pair-wise comparisons—between the reference level and each of the six experimental programs (3LF/2LF/1LF/1HF/2HF/3HF) and between conditions with the same magnitude of shift but in opposite directions (e.g., 3LF vs. 3HF)—were conducted using a Bonferroni-corrected α=.0055. Two- and three-channel shifted conditions were all significantly different from baseline [*p*<.0055], but one-channel shifts did not differ significantly from baseline. No significant differences were observed between condition pairs with the same magnitude shift in opposite directions. In summary, shifts of two and three channels (3 and 4.5 mm) in the poorer ear had a clear negative impact on bilateral scores, but shift direction did not influence this effect.

If the shifted signal in the poorer ear could be ignored, bilateral scores in the shifted conditions (open squares in Figs. 2 and 3) should have been no worse than unilateral better-ear alone scores (star symbols with dashed line in Figs. 2 and 3). This was not the case; bilateral scores in the two- and three-channel shifted conditions were lower, on average, than unilateral better-ear scores. A LMEM with experimental program treated as a categorical variable (6 shifted programs in the bilateral condition plus the unilateral better-ear condition as the reference), showed a main effect of experimental program [*F*(6,54)=13.66, *p*<.001]. Six pair-wise comparisons between the unilateral better-ear score and bilateral scores for each experimental program (3LF/2LF/1LF/1HF/2HF/3HF) were conducted using a Bonferroni-corrected α=.008. Scores in both three-channel shifted conditions (3LF and 3HF) were significantly lower than unilateral better-ear scores (*p*<.008) by about 15% (3LF: 14.4%, 3HF: 15.1%), but the differences for one- and two-channel shifts did not reach significance. Thus, at least for 3-channel (4.5-mm) shifts in the poorer ear, bilateral scores were poorer than unilateral better-ear scores, suggesting across-ear interference and an inability to ignore the shifted signal.

Although bilateral scores were the primary focus, the effects of frequency shift on unilateral scores were also of interest [open circles in Fig. 3(B)]. Previous vocoder research suggests that frequency shifts towards the base are more detrimental to unilateral sentence scores than equivalent shifts in millimeters towards the apex (e.g., Yoon *et al*., 2011). Additionally, although vowel recognition by unilateral CI users is minimally affected by frequency shifts <3 mm in either direction (Fu and Shannon, 1999), the direction and degree of shift might differently impact perception of full sentences. A LMEM analysis of unilateral poorer-ear scores was conducted with shift direction (base/apex; categorical) and degree of shift (1/2/3; ordered discrete values) included as fixed effects together with their interaction. Although a main effect of shift was observed [*F*(1,45)=108.6, *p*<.001], neither the main effect of direction (*p*=.35) nor the interaction (*p*=.98) were significant. Thus, shifts toward the base were not more detrimental than apical shifts. Additionally, pairwise comparisons showed that all levels of shift, including 1.5 mm, resulted in significantly lower unilateral scores than the baseline condition (corrected α=.017).

After the testing reported above, eight of the nine participants wore the shifted 3HF poorer-ear program and baseline better-ear program for one hour during informal social interaction. When this group was then retested in this bilateral 3HF Shift condition, there was a significant decrease in interference effect size [9.3% compared to 16.2% previously; *F*(1,8)=6.03, *p*=.04]. Bilateral scores nevertheless remained significantly lower than pre-training scores with the better-ear alone [*F*(1,8)=6.99, *p*=.03].

## 4. DISCUSSION

This study examined whether experimentally introduced interaural frequency mismatch influences sentence recognition in quiet for BI-CI participants. We found that shifts >3 mm reduced bilateral scores to levels poorer than for the better ear alone. Listeners were unable to attend only to the signal in their better ear when a distorted and frequency-shifted copy of the sentence was presented to their poorer ear.

The question of whether having a relatively poor input on one side can sometimes be disruptive to bilateral listening, debated in the hearing-aid literature (Jerger and Silverman, 2018), has also been raised for BI-CIs. Most previous research has explored the issue using BI-CI simulations. Yoon *et al*. (2011), for example, simulated interaural frequency mismatch by presenting normal-hearing listeners with six-channel sine-vocoded sentences. When frequency allocations were matched between ears in a trained baseline condition, bilateral scores were better than for a single ear alone. In contrast, with a 3-mm frequency shift towards the apex in one ear, bilateral scores were no better than for the baseline unshifted single ear alone. Finally, with a 3-mm frequency shift towards the base, bilateral scores were significantly worse than for the baseline unshifted single ear alone. The present BI-CI data are largely consistent with such vocoder studies, although shifts towards the apex were found to also be disruptive to bilateral listening.

The ability to attend to one ear and ignore the other is influenced by the absolute and relative spectral-temporal characteristics of the two signals (Goupell *et al*., 2021). Further research would be needed to better determine why listeners found it difficult to ignore the shifted signal in the present study. It is unknown if this result is specific to the frequency shift distortion used here or if other types of distortion might yield similar results. It is also worth noting that listeners were not explicitly told to ignore or attend to a particular ear. Instead, it was assumed that listeners would adopt the strategy that would most benefit task performance.

Frequency shift has been reported to hinder integration of bilateral signals even when using both signals is the optimal strategy. Using six-channel vocoded speech and alternate channels presented to each ear (even-numbered to the left, odd-numbed to the right) Siciliano *et al*. (2010) showed that after ten hours of training, normal-hearing listeners still could not integrate a 6-mm shifted signal in one ear with an unshifted complementary signal in the other ear to improve sentence scores over performance for the unshifted ear alone. Even though there was important, albeit highly distorted, speech information in the shifted ear, the normal-hearing participants did not use it to improve their speech recognition.

The present data are consistent with the idea that the degree of similarity between bilateral signals is critical to how the auditory system integrates or segregates inputs. Integrating information across two CIs offering dissimilar levels of benefit appears difficult and may play a role in how well binaural cues can be used to separate out target speech from non-target input (Goupell *et al*., 2018a). The 4.5-mm shifts in this study are outside the range of ±3 mm associated with monopolar electrical stimulation, beyond which significant decreases in BI-CI binaural processing occur (Goupell *et al*., 2022) and therefore should be large enough to hinder signal integration. It is worth noting, however that recent vocoder research suggests that with background noise or competing talkers, even small interaural frequency mismatches of 1-2 mm might reduce performance for BI-CI listeners (Xu *et al*., 2020).

The locus of the interference from interaural frequency mismatch is likely in the central auditory pathway. The reduced effect size after further exposure suggests that listeners can learn to partially reduce this interference, perhaps by focusing attention on only the better ear, although it should be noted that no control group was included to rule out possible effects of repeated testing. Additional testing with novel sentences or talkers would be useful to determine the type of learning taking place.

These results may contribute towards an understanding of why less-than-expected benefit from having two CIs is sometimes observed in BI-CI users. If large interaural frequency mismatch can impact speech perception even in quiet, appropriate clinical approaches to real-world cases might include programming adjustments guided by such measures as computed-tomography estimates of electrode position (Bernstein *et al*., 2021) to minimize such mismatch.

## Data Availability

All data produced in the present study are available upon reasonable request to the authors.

## Acknowledgments

This research was supported by the National Institute on Deafness and Other Communication Disorders of the National Institutes of Health under Award Number R01DC015798 (M.J.G., J.G.W.B.). The content is solely the responsibility of the authors and does not necessarily represent the official views or policy of the National Institutes of Health, the Department of Army/Navy/Air Force, Department of Defense, or US Government. *This article has been submitted to JASA Express Letters. If it is published it will be found at* http://asa.scitation.org/journal/jel

## Notes

### Competing Interest Statement

The authors have declared no competing interest.

### Author Declarations

The Institutional Review Board at University of Maryland College Park gave ethical approval for this work.

